# An optimized reference tissue method for quantification of tau protein depositions in diverse neurodegenerative disorders by PET with ^18^F-PM-PBB3 (^18^F-APN-1607)

**DOI:** 10.1101/2022.02.13.22270135

**Authors:** Kenji Tagai, Yoko Ikoma, Hironobu Endo, Oiendrila Bhowmik Debnath, Chie Seki, Kiwamu Matsuoka, Hideki Matsumoto, Masaki Oya, Kosei Hirata, Hitoshi Shinotoh, Keisuke Takahata, Shin Kurose, Yasunori Sano, Maiko Ono, Hitoshi Shimada, Kazunori Kawamura, Ming-Rong Zhang, Yuhei Takado, Makoto Higuchi

**Affiliations:** Institute for Quantum Medical Science, Quantum Life and Medical Science Directorate, National Institutes for Quantum and Radiological Science and Technology, Chiba 263-8555, Japan; Department of Psychiatry, The Jikei University of Medicine, Tokyo 105-8461, Japan; Department of Psychiatry, Keio University School of Medicine, Tokyo 160-0016, Japan; Department of Functional Neurology & Neurosurgery, Center for Integrated Human Brain Science, Brain Research Institute, Niigata University, Niigata 951-8585, Japan

**Author notes:** These authors contributed equally (Kenji Tagai, Yoko Ikoma).

**Keywords:** tau PET, reference tissues, Alzheimer’s disease, progressive supranuclear palsy, frontotemporal lobar degeneration

## Abstract

Positron emission tomography (PET) with ^18^F-PM-PBB3 (^18^F-APN-1607) enables high-contrast detection of tau depositions in various neurodegenerative dementias, including Alzheimer’s disease (AD) and frontotemporal lobar degeneration (FTLD). A simplified method for quantifying the radioligand binding in target regions is to employ the cerebellum as a reference (CB-ref) on the assumption that the cerebellum has minimal tau pathologies. This procedure could be valid in AD, while FTLD disorders exemplified by progressive supranuclear palsy (PSP) are characterized by occasional tau accumulations in the cerebellum, hampering the application of CB-ref. The present study aimed to establish an optimal method for defining reference tissues on ^18^F-PM-PBB3-PET images of the AD and non-AD tauopathy brains. We developed a new algorithm to extract reference voxels with a low likelihood of containing tau deposits from gray matter (GM-ref) or white matter (WM-ref) by a bimodal fit to an individual, voxel-wise histogram of the radioligand retentions and applied it to ^18^F-PM-PBB3-PET data obtained from age-matched 40 healthy controls (HCs) and 23 AD, 40 PSP, and five other tau-positive FTLD patients. PET images acquired at 90-110 min after injection were averaged and co-registered to corresponding magnetic resonance imaging space. Subsequently, we generated standardized uptake value ratio (SUVR) images estimated by CB-ref, GM-ref and WM-ref respectively, and then compared the diagnostic performances. GM-ref and WM-ref covered a broad area in HCs and free of voxels located in regions known to bear high tau burdens in AD and PSP patients. GM-ref allowed the most robust separation of AD and PSP patients from HCs according to the area under the curves in receiver operating characteristic curve analyses. GM-ref also provided SUVR images with higher contrast than CB-ref in FTLD patients with suspected and confirmed corticobasal degeneration. The methodology for determining reference tissues as optimized here reinforces the accuracy of ^18^F-PM-PBB3-PET measurements of tau burdens in a wide range of neurodegenerative illnesses.

## 1. Introduction

Depositions of tau fibrils in the brain are the hallmarks of Alzheimer’s disease (AD) and a significant subset of frontotemporal lobar degeneration (FTLD), including progressive supranuclear palsy (PSP), corticobasal degeneration (CBD), and Pick’s disease (PiD). The structures of tau in these and diverse other tauopathies are distinct (1, 2) and determine the subcellular, cellular, and regional distribution of tau aggregates in close relationships with clinical phenotypes (3, 4).

The recent development of radioligands for positron emission tomography (PET) has enabled *in vivo* visualization of tau fibril depositions (5, 6). To quantify tau accumulations in the AD brains, a ratio of the radioactivity uptake between target and reference tissue defined as standardized uptake value ratio (SUVR) or distribution volume ratio is often employed as a simplified index for the specific probe binding to tau aggregates under the assumption that the reference region is devoid of tau lesions harboring binding components. Similarly, time-radioactivity curves in the target and reference regions are comparatively utilized for the estimation of non-displaceable binding potential (BP_ND_) of the radiotracer according to pharmacokinetic models. The reference tissue is primarily defined on the cerebellar gray matter (CB-ref) for PET measurements of AD-type tau pathologies since this anatomical structure is supposed to contain minimal tau fibrils in AD (7-10).

While most PET probes for tau deposits are incapable of sensitive detection of FTLD-type tau assemblies formed by a subgroup of tau isoforms, we developed a PET ligand, ^11^C-PBB3, for visualizing AD and non-AD tau pathologies, including PSP-, CBD-, and PiD-type tau inclusions (11). ^11^C-PBB3 did not yield abundant radiosignals in the brain due to its high propensity to metabolic conversions and consequent inefficiency of its transfer to the brain, but a fluorinated analog of ^11^C-PBB3, ^18^F-PM-PBB3 (also known as ^18^F-APN-1607), displayed improved biostability and allowed detection of a wide range of tau lesions with high contrast (12). The use of CB-ref might lead to underestimation of the radiotracer binding in target tissues, particularly in patients with PSP and CBD, as tau pathologies could exist in the cerebellum of these cases (13, 14). To circumvent this technical issue, we extracted reference voxels with a low likelihood of tau depositions from gray matter in parametric images based on whether the voxel-wise BP_ND_ was within a range assigned using a histogram in healthy controls (13). This method was applied to determining BP_ND_ of ^11^C-PBB3 in subjects with AD (13), PSP (14), and several other non-AD tauopathies (15, 16). More recently, other research groups constituted a procedure to extract reference voxels from white matter based on whether the voxel-wise radiotracer retention was included in a range assigned using an individual histogram (17, 18). This methodology was then implemented in the estimation of ^18^F-PM-PBB3 retention in AD cases (18), and the employment of individual histograms allowed handy quantitative assays without generating an average histogram in control subjects. However, it remains unclear whether reference voxels should be collected from gray or whiter matter, and how the range of the probe retention in the histogram is selected for the reference extraction also needs to be determined.

The present study aims to establish an optimal method to define reference voxels for the quantification of ^18^F-PM-PBB3 binding in diverse tauopathies. We constructed a new workflow to assign the range of the probe uptake for picking up adequate voxels with sufficient robustness in consideration of diverse histogram profiles. Then, the accuracy of diagnostic discriminations and contrasts for tau pathologies were compared among CB-ref and pools of reference voxels extracted from gray matter (GM-ref) and white matter (WM-ref).

## 2. Methods

### 2.1. Participants

We analyzed age-matched datasets of 40 healthy controls (HCs), 23 patients with AD, 40 patients with PSP, and five patients with other FTLD syndromes. These datasets were acquired from clinical studies registered in UMIN Clinical Trials Registry (UMIN-CTR; IDs 000030248, 000034546, 000033808, and 000030319). They were approved by the Radiation Drug Safety Committee and National Institutes for Quantum Science and Technology Certified Review Board of Japan. Written informed consent was obtained from all subjects and/or from close family members when subjects were cognitively impaired.

All patients were clinically diagnosed according to the established criteria as we previously reported (19-24). Depositions of amyloid-beta (Aβ) were assessed by a visual inspection of ^11^C-PiB-PET images (12). All AD patients were indicated by PET to have Aβ plaques and denoted as Aβ (+), and five Aβ (+) MCI patients were added to the AD group. The PSP group consisted of 33 patients with PSP Richardson syndrome (PSP-Richardson) with typical manifestations and seven patients with other clinical phenotypes (PSP-other). HCs were without a history of neurological and psychiatric disorders, and they were age-matched with the AD and PSP groups (Table 1). Patients with other FTLDs, including corticobasal degeneration syndrome (CBS), progressive non-fluent aphasia (PNFA), and behavioral variant of frontotemporal dementia (BvFTD), were also incorporated as in our previous report (12), and two of these cases were neuropathologically diagnosed as having CBD and PiD by biopsy and autopsy, respectively (25). All HCs, PSP, and other FTLD patients were Aβ(-) according to PET findings.

**Table 1:**
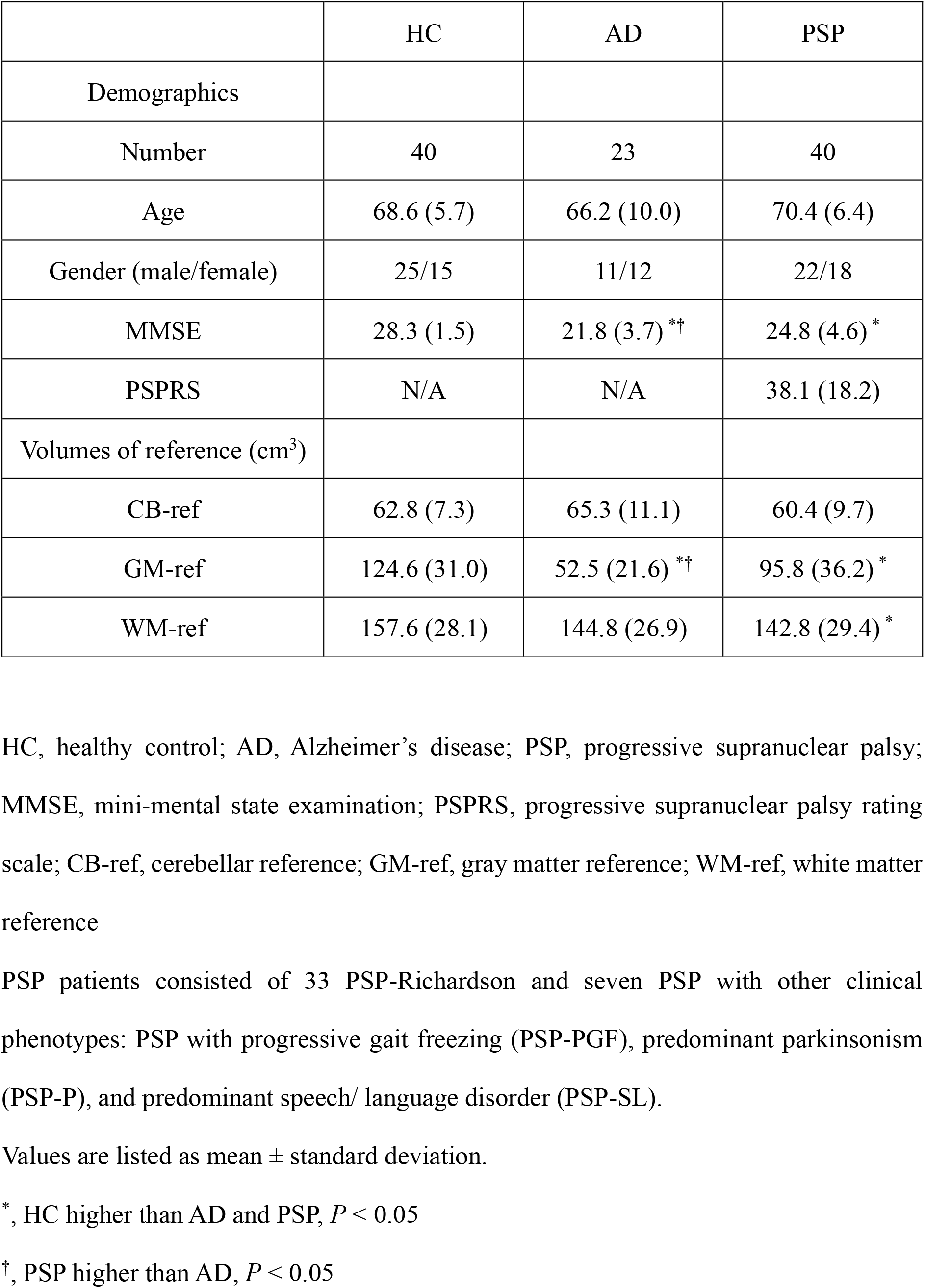
Demographics and characteristics of each reference.

|  | HC | AD | PSP |
| --- | --- | --- | --- |
| Demographics |  |  |  |
| Number | 40 | 23 | 40 |
| Age | 68.6 (5.7) | 66.2 (10.0) | 70.4 (6.4) |
| Gender (male/female) | 25/15 | 11/12 | 22/18 |
| MMSE | 28.3 (1.5) | 21.8 (3.7) <sup>*†</sup> | 24.8 (4.6) <sup>*</sup> |
| PSPRS | N/A | N/A | 38.1 (18.2) |
| Volumes of reference (cm<sup>3</sup>) |  |  |  |
| CB-ref | 62.8 (7.3) | 65.3 (11.1) | 60.4 (9.7) |
| GM-ref | 124.6 (31.0) | 52.5 (21.6) <sup>*†</sup> | 95.8 (36.2) <sup>*</sup> |
| WM-ref | 157.6 (28.1) | 144.8 (26.9) | 142.8 (29.4) <sup>*</sup> |

HC, healthy control; AD, Alzheimer's disease; PSP, progressive supranuclear palsy; MMSE, mini-mental state examination; PSPRS, progressive supranuclear palsy rating scale; CB-ref, cerebellar reference; GM-ref, gray matter reference; WM-ref, white matter reference
PSP patients consisted of 33 PSP-Richardson and seven PSP with other clinical phenotypes: PSP with progressive gait freezing (PSP-PGF), predominant parkinsonism (PSP-P), and predominant speech/ language disorder (PSP-SL).
Values are listed as mean ± standard deviation.
<sup>\*</sup>, HC higher than AD and PSP, $P < 0.05$
<sup>†</sup>, PSP higher than AD, $P < 0.05$

### 2.2. Image acquisition and data preprocessing

MR images were acquired with a 3-T scanner, MAGNETOM Verio (Siemens Healthcare). Three-dimensional T1-weighted gradient-echo sequence produced a gapless series of thin sagittal sections (TE = 1.95 ms, TR = 2300 ms, TI = 900 ms, flip angle = 9°, acquisition matrix = 512×512×176, voxel size = 1 × 0.488 × 0.488 mm). PET assays were conducted with a Biograph mCT Flow system (Siemens Healthcare), which provides 109 sections with an axial field of view of 16.2 cm. The intrinsic spatial resolution was 5.9 mm in-plane and 5.5 mm full-width at half-maximum axially. Images were reconstructed using a filtered back-projection algorithm with a Hanning filter (4.0 mm full-width at half-maximum). ^18^F-PM-PBB3 were injected into the subjects with an average dose of 186.4 ± 8.2 MBq and a molar activity of 237.4 ± 75.3 GBq/μmol.

Data preprocessing was performed using PMOD 3.8 (PMOD Technologies Ltd) and Statistical Parametric Mapping software (SPM12, Wellcome Department of Cognitive Neurology). Acquired PET images were corrected for head motions and then rigidly co-registered to individual T1-weighted MR images. To generate standardized uptake value ratio (SUVR) images, we averaged PET data acquired at 90 - 110 min after radiotracer injection. In addition, the individual MR images were segmented, and the probability maps of gray matter (GM) and white matter (WM) were generated using SPM12 for the extraction of reference voxels based on histogram analysis.

### 2.3. Histogram-based definition of reference voxels

We propose a new method to define reference voxels in GM or WM based on the frequency histogram of voxel-counts with a homemade script implemented MATLAB (The Mathworks, Natick, MA, USA). A binary mask image of GM or WM was generated from the probability maps by selecting voxels with a higher than 90% probability of being GM or WM. The erosion with morphological operation using 3 × 3 × 3 neighboring voxels was also conducted on each mask to eliminate influence from the boundary regions. Subsequently, a voxel-wise frequency histogram of voxel-counts was constructed from an individual PET image masked for GM or WM.

A bimodal Gaussian distribution fit was then applied to the histogram in light of the view that voxels with and without tau pathologies formed two distinct peaks. The first Gaussian distribution with the lower-count peak was regarded to contain voxels with no or minimal tau deposits, and hence voxels with values within the full-width at half-maximum (FWHM) of this peak were extracted and pooled as a reference region. Tracer retention in the extracted reference region (*C*_ref_) was determined by averaging reference voxel values weighted for the probability of being contained in the first versus second Gaussian distribution (*w*_i_) as follows:

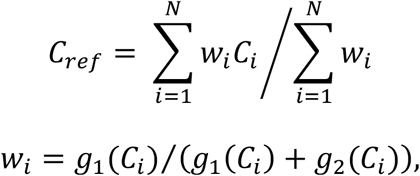

where *C*_i_ is the value of the i’th voxel, *g*1 and *g*2 are the first and second Gaussian distribution functions, respectively, and *N* is the number of reference voxels (Figure 1A). If the height of the first peak was less than half of the second peak, we applied monomodal Gaussian distribution to the histogram to circumvent potential lack of the fitting robustness in the bimodal fit due to an insufficient number of voxels in the first peak (Figure 1B). The monomodal Gaussian distribution fit was then evaluated by the Dice coefficient, which is a ratio of the area between the doubled intersection of the polygonized histogram and fitted Gaussian distribution and the sum of these two polygons. If the Dice coefficient was smaller than the threshold value determined with HC data in the current cohort, the bimodal Gaussian distribution fit was considered more appropriate than the monomodal fit (Figure 1C). The threshold was set at 0.936, which is the mean - 2 standard deviation (SD) value of cases in which monomodal fitting was applied in HCs. In the use of the monomodal fit, *C*_ref_ was determined by averaging counts in all voxels within the FWHM (Figure 1B). Finally, all voxel-counts were normalized by *C*_ref_ in GM-ref or WM-ref as well as retention in CB-ref (cerebellar cortex) labeled with Freesurfer 6.0 from the Desikan–Killiany–Tourville atlas as a conventional method (25), to produce SUVR images, respectively.

**Figure 1:**
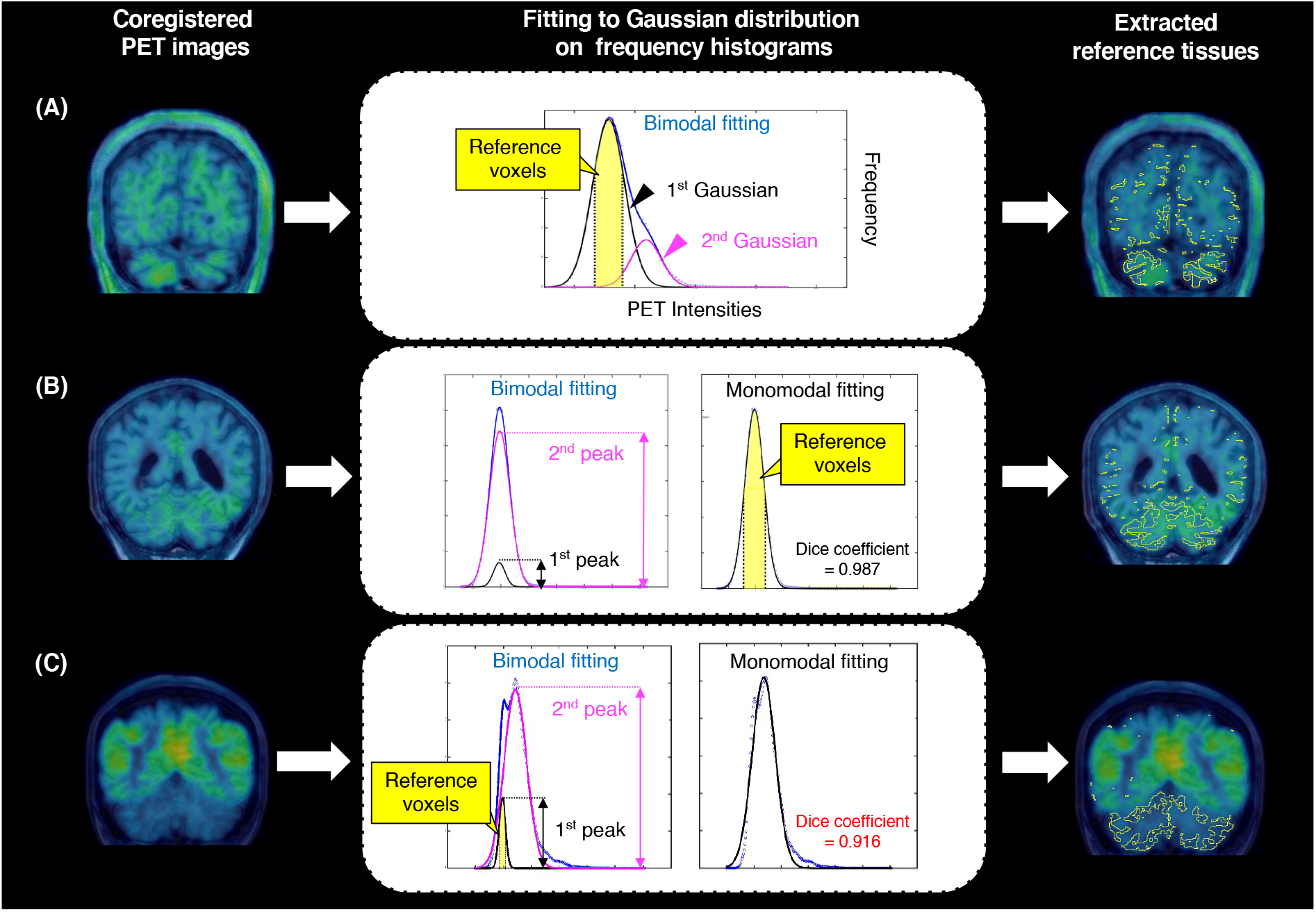
Flowcharts of extracting reference tissues by developed algorithm on ^18^F-PM-PBB3 PET images. The yellow-labeled regions show extracted references based on bimodal/monomodal Gaussian fitting, respectively. (A) A histogram obtained from a PSP-Richardson case, and the reference was extracted from the 1st Gaussian distribution based on bimodal fitting. (B) shows histograms obtained from a PSP-PGF case. The amplitude of the 1st peak was less than half of the 2nd peak on bimodal fitting (left), and the Dice coefficient was higher than the threshold value (>0.939) on monomodal fitting (right). Hence, the reference was set based on monomodal fitting. (C) shows histograms obtained from an AD case. The amplitude of the 1st peak was less than half of the 2nd peak on bimodal fitting (left), whereas the Dice coefficient was lower than the threshold value (<0.939) on monomodal fitting (right). Ultimately, the reference was set based on bimodal fitting.

### 2.4. Target regions

The target region was placed at an area with abundant tracer binding in AD and PSP patients, according to our previous studies (12). We defined volumes of interest (VOIs) in the neocortex involved in AD tau pathologies at the Braak stages V and VI (BraakV/VI) and in the subthalamic nucleus (STN) burdened with PSP-type tau deposits. The Braak V/VI VOI was labeled using FreeSurfer 6.0 as described elsewhere. The STN VOI was defined with a template atlas (Talairach Daemon atlas from the Wake Forest University PickAtlas version 3.0.5) in the MNI (Montreal Neurologic Institute) space, and spatial normalization was conducted according to the Diffeomorphic Anatomical Registration Through Exponentiated Lie Algebra (DARTEL) algorithm. For other FTLDs, VOIs were also generated with FreeSurfer 6.0 in areas enriched with tau lesions characteristic of each disease, such as the precentral cortex in CBS and PNFA and the orbitofrontal cortex in BvFTD.

### 2.5. Statistical analyses

Statistical examinations were performed using GraphPad Prism 9.0. We adapted Fisher’s exact test (sex) and Kruskal-Wallis test (other parameters) for comparisons between HCs, AD, and PSP groups (*P* < 0.05, corrected for Dunn’s multiple comparisons). The performance of the diagnosis based on SUVR values in the target VOIs against CB-ref, GM-ref, and WM-ref was evaluated using Mann-Whitney U test and area under the curve (AUC) values in Receiver Operating Characteristic (ROC) curves. Subsequently, linear regression analysis was conducted to assess correlations between SUVRs estimated with these three reference tissues.

## 3. Results

### 3.1. Evaluations of reference tissues

Table 1 shows the demography of the subjects and volumes of defined CB-ref, GM-ref, and WM-ref. The bimodal Gaussian distribution fit to the histogram was judged as adequate in most cases, while the monomodal fit was chosen in GM histograms of six HCs and five PSP, and WM histograms of one HC and one AD case. Figure 2 demonstrates representative images of GM-ref and WM-ref. As a result of Gaussian fitting, voxels in GM-ref showed a tendency to pick up from the entire cortex in HCs, the cerebellum in AD, and regions excluding the cerebellum in PSP cases. Meanwhile, voxels in WM-ref were extracted from the entire WM in HCs and AD and regions excluding a WM portion of the basal ganglia in PSP.

**Figure 2:**
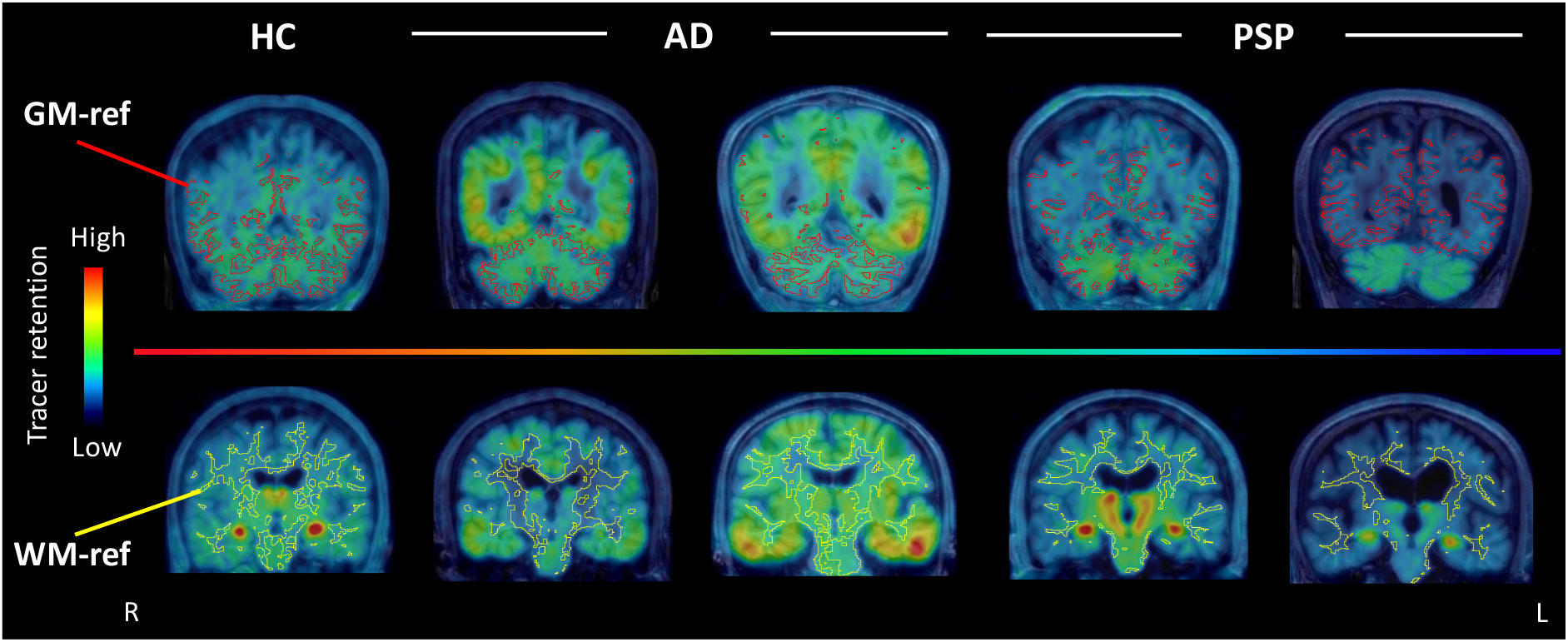
Comparisons of characteristics of GM-ref and WM-ref generated using histograms. References generated by the histograms in representative cases of HCs and patients with AD and PSP. All PET images were co-registered T1-weighted MR images; red labeled regions demonstrate generated GM-ref (upper row); yellow labeled regions show WM-ref (lower row).

### 3.2. Diagnostic performance of quantifications with CB-ref, GM-ref, and WM-ref

Figure 3 illustrates comparisons of SUVRs in the target VOIs among the AD, PSP, and HC groups and ROC curve analyses for the discrimination of AD or PSP patients from HCs. SUVRs determined with all three reference tissues exhibited significant differences between the patient and HC groups (*p* < 0.05, Figure 3A, B), whereas the SD of these values estimated with WM-ref was higher than those estimated with CB-ref and GM-ref in HCs (Supplemental Table 1).

**Figure 3:**
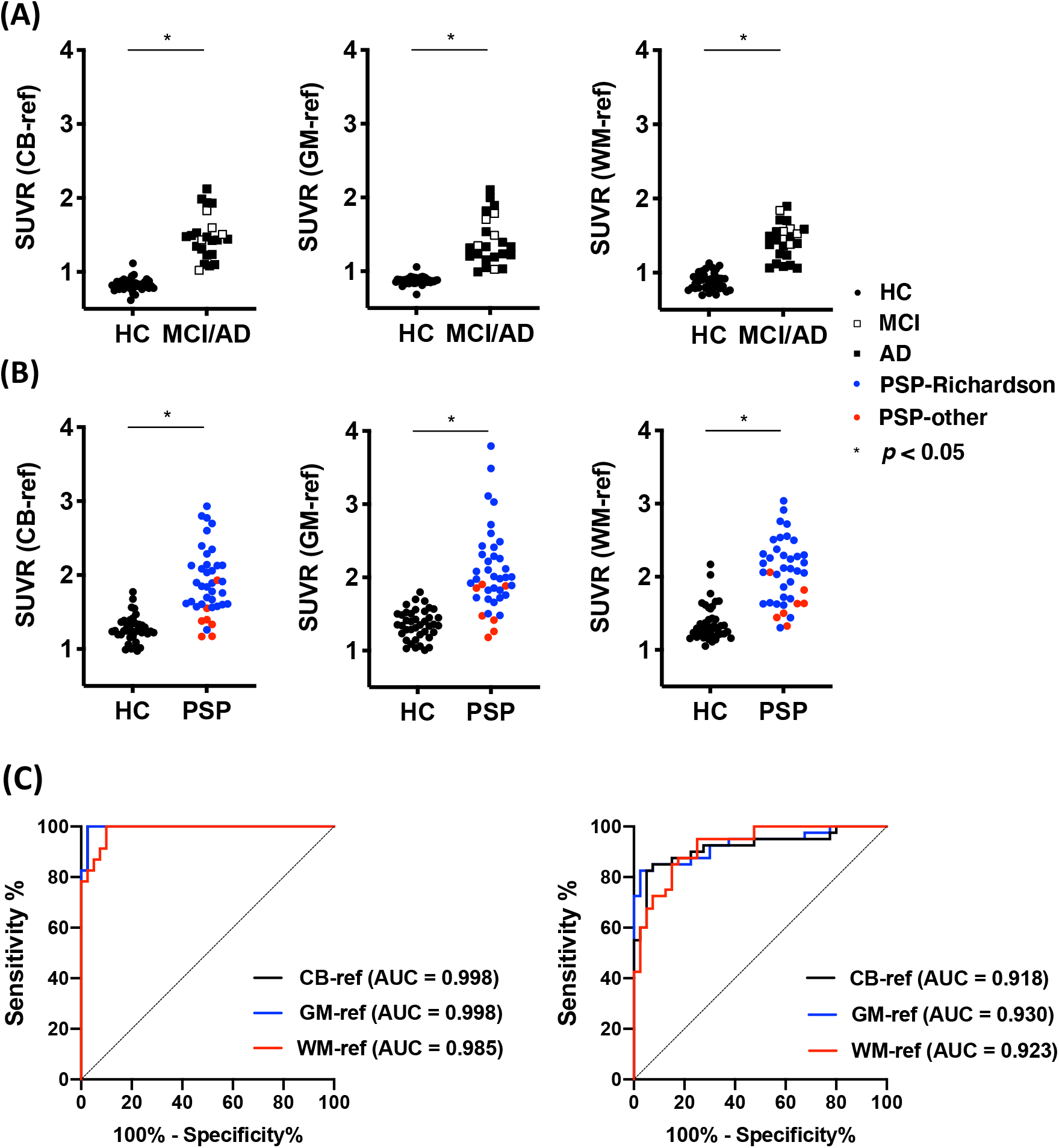
Comparison of diagnostic performances for discriminating patients from HCs between each reference. (A, B) Scatter plots show SUVR values of Braak V/VI (A) and STN (B) in the case of quantified by each reference. The color of each dot indicates the clinical profile: Black circles; (HC), white squares; (MCI), black squares; (AD), blue circles; (PSP-Richardson) and red circles; (PSP-other). Asterisks represent P < 0.05 by Mann-Whitney U test. (C) ROC curves estimated from each reference are shown for the following groups: HCs vs MCI/AD (left) and PSP (right). Each color shows the type of references: black (CB-ref), blue (GM-ref), and red (WM-ref).

In ROC curve analyses, the use of CB-ref and GM-ref yielded high AUC values (= 0.998) relative to WM-ref (= 0.985) in the separation between AD patients and HCs (Figure 3C). GM-ref also produced a higher AUC value (= 0.930) than WM-ref (= 0.923) and CB-ref (= 0.918) in the discrimination between PSP patients and HCs (Figure 3C).

### 3.3. Comparisons of imaging data yielded by CB-ref and GM-ref

We then compared parametric SUVR images generated with CB-ref and GM-ref as these reference tissues enabled better differentiation between the patients and HCs than WM-ref. The analyses with CB-ref and GM-ref similarly captured spreading of the tracer retention from the medial and inferior temporal regions to the other neocortical areas along with cognitive declines in the continuum from MCI to AD (Figure 4A). Moreover, SUVRs in the target VOIs estimated with GM-ref and CB-ref were in good agreement with each other in MCI and AD cases (Figure 4B). By contrast, the quantification using GM-ref detected tau depositions in PSP patients with higher SUVR than the analysis with CB-ref (Figure 5A). Notably, tau accumulations primarily in the midbrain and left neocortical areas were undetectable with CB-ref but were clearly visualized with GM-ref in a PSP patient with aphasia (PSP-SL) (Figure 5A). GM-ref yielded higher SUVR values in the target VOIs than CB-ref in these PSP cases, indicating high contrast for tau aggregates provided by GM-ref (Figure 5B).

**Figure 4:**
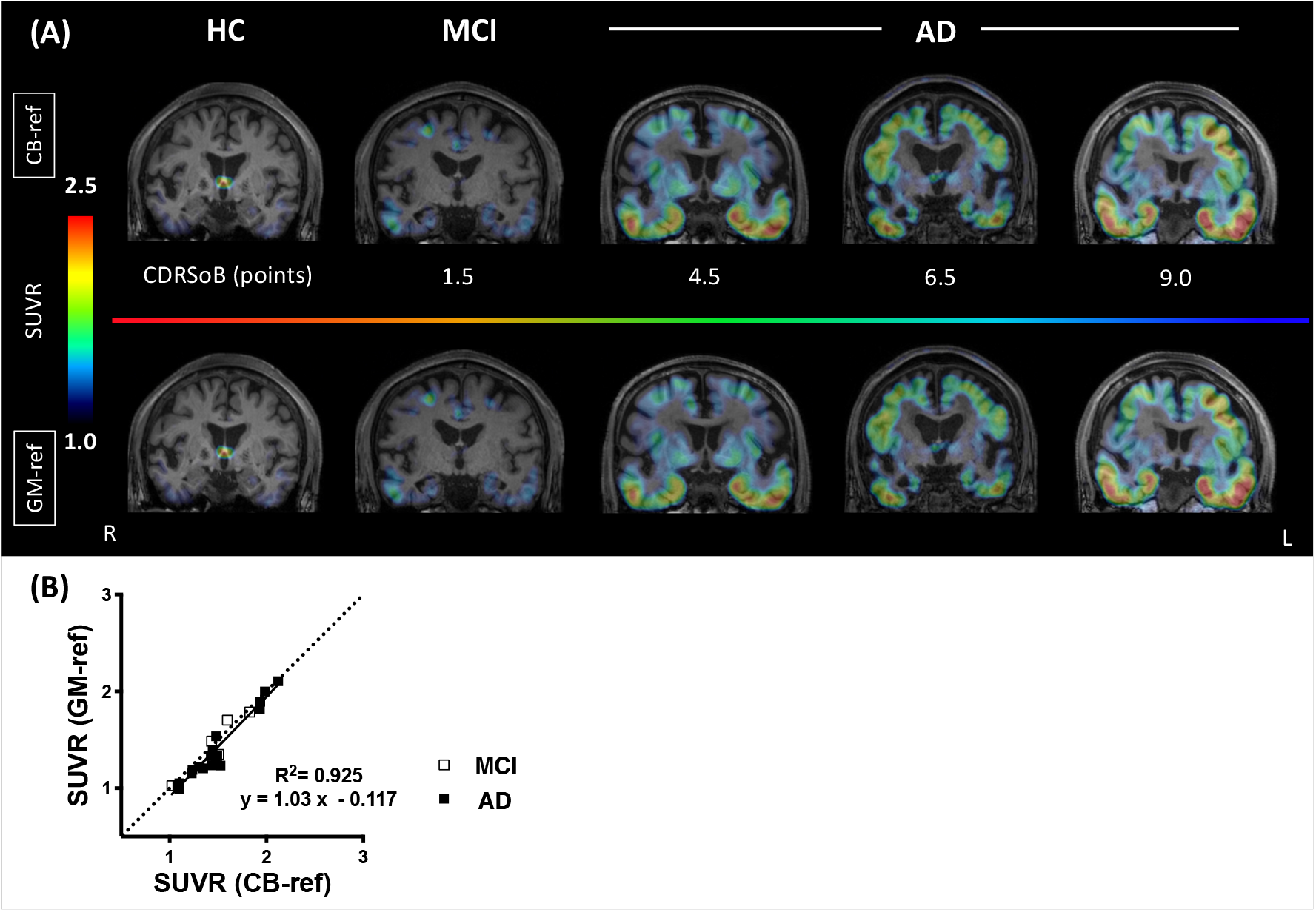
Direct comparison of SUVRs quantified by CB-ref and GM-ref in HCs and AD patients. (A) SUVR parametric images quantified by CB-ref in the upper row and by GM-ref in the lower row are shown. Tau lesions are visualized with similar contrast to CB-ref from MCI to AD. (B) The Scatterplot demonstrates a linear regression analysis. Black circles: (HC), white squares: (MCI), black squares: (AD). SUVR values are estimated from BraakV/VI VOI.

**Figure 5:**
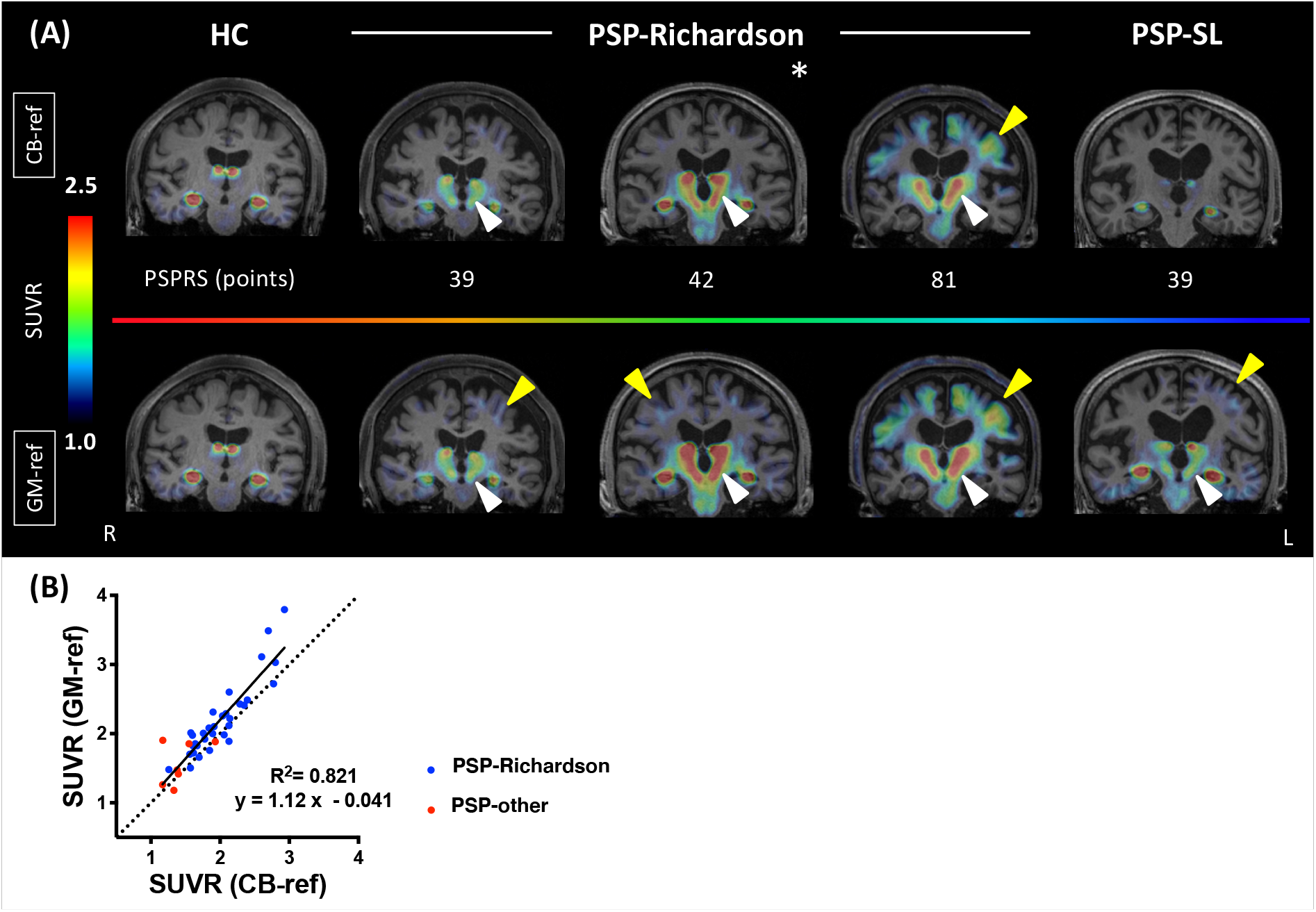
Direct comparison of SUVRs quantified by CB-ref and GM-ref in HCs and PSP patients. (A) SUVR parametric images quantified by CB-ref in the upper row and by GM-ref in the lower row are shown. Tracer retention was increased and visualized in GM-ref compared to CB-ref in areas with well-known PSP pathology such as basal ganglia (white arrowheads) and motor cortex (yellow arrowheads). The asterisk image was obtained from a neuropathologically confirmed PSP patient. (B) The Scatterplot demonstrates a linear regression analysis. Blue circles, (PSP-Richardson); red circles, (PSP with other clinical phenotypes). SUVR values are estimated from STN VOI.

We also assessed individual SUVR images of patients with various clinical and/or neuropathological phenotypes of non-PSP FTLDs (Figure 6). In a case with neuropathologically confirmed CBD, the assay with GM-ref captured tau depositions in the motor cortex and subcortical areas more sensitively than the use of CB-ref. Besides, GM-ref allowed imaging of tau pathologies with topologies characteristic of PSP and CBD in Non-AD CBS and PNFA patients with higher contrast than CB-ref. Finally, SUVR images generated with CB-ref and GM-ref displayed high similarity in BvFTD patients with suspected or confirmed PiD neuropathology, presumably due to the lack of tau aggregates in the cerebellum.

**Figure 6:**
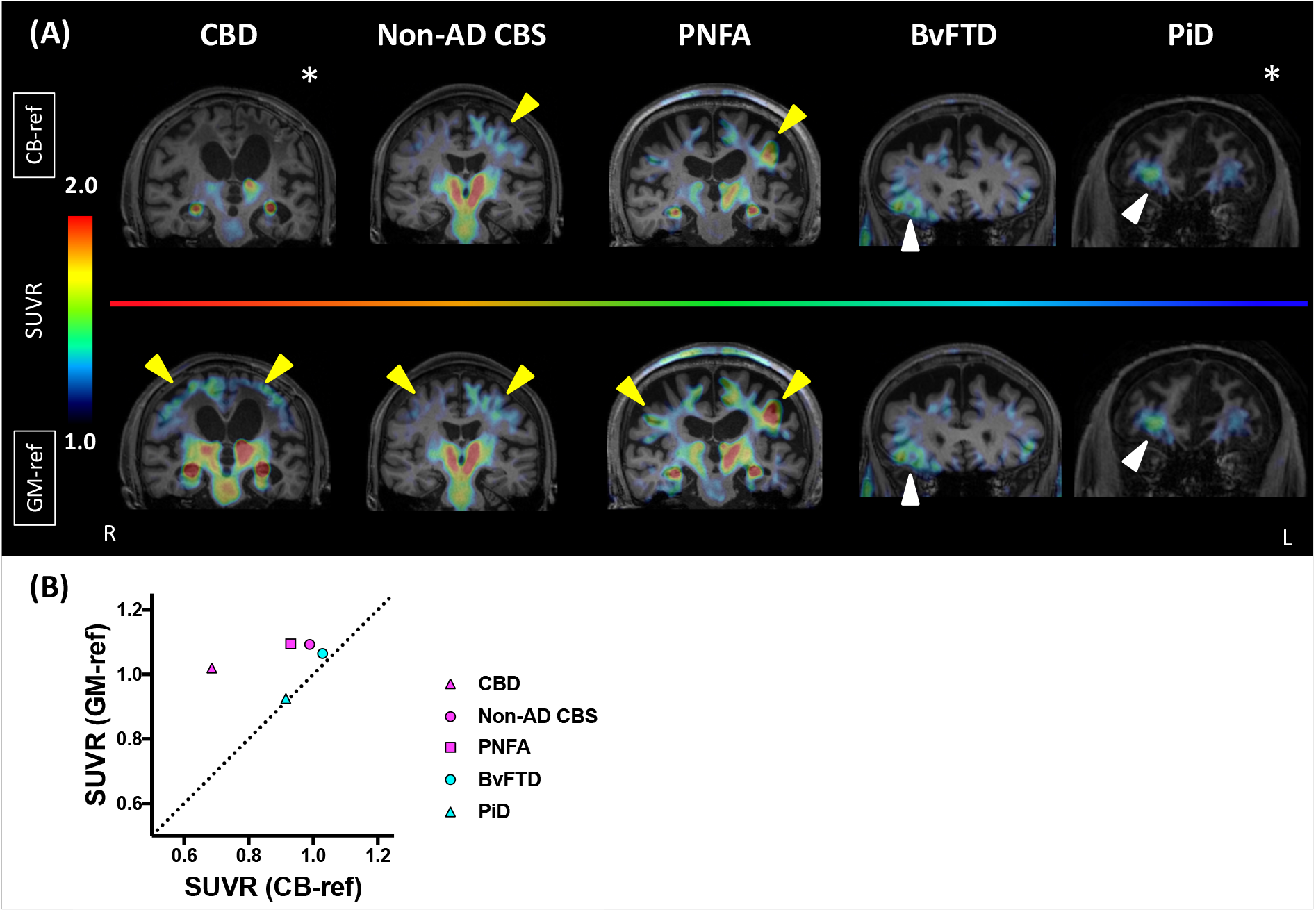
Direct comparison of SUVRs quantified by CB-ref and GM-ref in other FTLD patients. (A) SUVR parametric images quantified by CB-ref in upper row and by GM-ref in lower row are shown. Tracer retention was increased and visualized in GM-ref compared to CB-ref in CBD, Non-AD CBS, PNFA and patients but remained almost the same in BvFTD and PiD patients. Arrowheads indicate each target region: motor (yellow) and orbito (white) frontal cortices. Asterisked images were derived from neuropathologically confirmed patients. (B) Comparison of SUVR values quantified by CB-ref and GM-ref n each target region. Each target region was set at precentral cortex of CBD (magenta triangle), Non-AD CBS (magenta circle) and PNFA (magenta square) patients, and orbitofrontal cortex of BvFTD (light blue circle) and PiD (light blue triangle).

## 4. Discussion

In the present work, we have optimized the determination of reference tissue for the sensitive and precise PET detection of tau pathologies in various neurodegenerative dementias. This methodology reinforces the advantage of ^18^F-PM-PBB3 as an imaging agent for AD and non-AD tau depositions. In addition to the conventional CB-ref placement, Gaussian fits to individual histograms of ^18^F-PM-PBB3 retentions in GM and WM segments provided a range of radioligand uptakes for the extraction of voxels with a low likelihood of possessing tau deposits. Notably, GM-ref performed as accurately as CB-ref and better than WM-ref in the discrimination between AD-spectrum cases and HCs and enabled more robust differentiation of PSP patients from HCs than did CB-ref and WM-ref. Accordingly, our findings rationalize the utilization of GM-ref for capturing and quantifying different types of tau accumulations in any of the brain regions, including the cerebellum.

The application of histograms enabled us to exclude regions that were likely to contain specific binding and to extract optimized reference regions regardless of the type of diseases (13, 17). A vital issue in the histogram-based definition of reference voxels is the choice of an adequate mathematical model to describe the observed curves, as exemplified by monomodal and bimodal Gaussian fits. In the employment of our precedent tau PET probe, ^11^C-PBB3, a relatively low dynamic range for the detection of specific binding components impeded the application of a bimodal fit to the acquired image data (11, 13). By contrast, high contrasts for tau deposits produced by ^18^F-PM-PBB3 have allowed clear separation between two histogram clusters representing voxels with and without noticeable tau pathologies and voxels lacking PET-visible tau fibrils constitute the first, larger peak (Fig. 1A). Meanwhile, the two peaks can be barely discriminated in histograms of HCs burdened with few tau aggregates and PSP cases harboring low-grade tau depositions, justifying the use of a monomodal fit in these subjects (Fig. 1B). Moreover, the first peak may be small but distinguishable from the second peak in patients with advanced AD, and GM-ref extracted by the bimodal fitting resembled CB-ref (Fig. 1C).

An additional advantage of the current procedure over the previous method (13) is the assignment of a radioligand uptake range for the selection of reference voxels in each individual without introducing average cutoff values determined in the HC group. The group-wise cutoff could vary in a manner dependent on PET scanners and image reconstruction algorithms, precluding the unification of tau measurements among different PET facilities. The circumvention of this drawback in the present workflow will therefore facilitate multicenter tau PET assessments of elderly subjects on a large scale. In this study, GM-ref yielded higher AUC values in the ROC analysis than WM-ref across the diseases, which was primarily attributed to the variability in the quantification of SUVRs with WM-ref in HCs. WM-ref had been reported to be valid in several amyloid and tau PET studies (17, 26, 27) as its large size could be advantageous for reducing the statistical variability relative to CB-ref. In the meantime, non-specific retentions of these radioprobes in WM areas have been observed to vary among HCs partially in relation to aging (28, 29). It is also noteworthy that even slight alterations of off-target tracer binding may lead to large variabilities of SUVRs in the use of WM-ref if the background (free) tracer retention in WM is very low. Although factors affecting the non-displaceable uptake of ^18^F-PM-PBB3 in WM are yet to be identified, we consider GM-ref to be preferable to WM-ref for the quantification of tau deposits with this radioligand.

GM-ref also exhibited better diagnostic performance than CB-ref in patients with FTLD disorders, particularly in those with putative and confirmed PSP and CBD pathologies. Indeed, PSP tau pathologies could involve the cerebellar dentate nucleus and adjacent WM from a relatively early stage (30, 31). Similarly, tau lesions can be found in these areas of atypical or advanced CBD cases (32-34). Furthermore, tau inclusions in cerebellar GM Purkinje cells have also been detected in both PSP and CBD patients (35, 36). Thus, the localization of these pathological tau aggregates could undermine the validity of CB-ref for the PET assessment of FTLD disorders. The histogram-based reference definition proposed here can exclude voxels with potential tau pathologies throughout the entire GM or WM, including the cerebellar regions, in an unbiased fashion. In fact, GM-ref was composed of extensive neocortical areas rather than the cerebellar sectors in consecutive PSP and CBD cases, along with CBS and PNFA patients suspected of having PSP or CBD pathologies, and low-grade tau accumulations in neocortical structures of these subjects could be captured by using GM-ref but not CB-ref. By contrast, tau depositions in PiD patients were nearly equally detectable by applying GM-ref and CB-ref, in agreement with the lack of intense cerebellar tau lesions in this disorder.

As mentioned above, GM-ref and CB-ref showed comparable diagnostic performance in discriminating AD cases from HCs. We also postulate that GM-ref can utilize a larger volume of brain areas than CB-ref in the assays of patients with prodromal and early AD, contributing to a gain of the quantification stability. This potential benefit would help sensitive detection of tau accumulations with low abundances in incipient AD.

In conclusion, the newly developed workflow for the determination of reference tissue fortifies the utility of ^18^F-PM-PBB3 for investigating a broad spectrum of neurodegenerative tauopathies with high contrast. The tau PET technology in conjunction with this quantitative procedure will also permit longitudinal measurements of tau aggregates with minimal influences of temporal changes in specific and non-displaceable binding components at a certain location, which is advantageous in the evaluation of emerging anti-tau therapeutics for AD and FTLD disorders.

## Data Availability

Requests for data that support the findings of this study should be directed to Correspondence, Kenji Tagai and Yuhei Takado, and will be available upon reasonable request.

## Credit authorship contribution statement

**Kenji Tagai, Yoko Ikoma:** Conceptualization, Methodology, Formal analysis, Writing - original draft. **Hironobu Endo, Oriendrila Debnath, Chie Seki, Maiko Ono**: Conceptualization, Methodology. **Kiwamu Matsuoka, Keisuke Takahata, Hitoshi Shinotoh, Hideki Matsumoto, Masaki Oya, Kosei Hirata, Shin Kurose, Yasunori Sano, Hitoshi Shimada**: Collection of clinical data. **Kazunori Kawamura, Ming-Rong Zhang**: Radioligand synthesis. **Yuhei Takado, Makoto Higuchi**: Conceptualization, Writing - review & editing, Funding acquisition, Supervision

## Acknowledgements

The authors thank all patients and their caregivers for participation in this study, as well as clinical research coordinators, PET and MRI operators, radiochemists, and research ethics advisers at QST for their assistance with the current projects. We thank APRINOIA Therapeutics for kindly sharing a precursor of ^18^F-PM-PBB3. The authors acknowledge support with the recruitment of patients by Shunichiro Shinagawa at the Department of Psychiatry, Jikei University School of Medicine; Shigeki Hirano at the Department of Neurology, Chiba University; Taku Hatano, Yumiko Motoi, and Shinji Saiki at the Department of Neurology, Juntendo University School of Medicine; Ikuko Aiba at the Department of Neurology, National Hospital Organization Higashinagoya National Hospital; Yasushi Shiio and Tomonari Seki at the Department of Neurology, Tokyo Teishin Hospital; Hisaomi Suzuki at the National Hospital Organization Shimofusa Psychiatric Medical Center.

## Supplementary materials

Supplementary material associated with this article can be found in the online version.

