## Supplemental Table 1 for "An optimized reference tissue method for quantification of tau protein depositions in diverse neurodegenerative disorders by PET with ^18^F-PM-PBB3 (^18^F-APN-1607)"

**Supplemental Table 1: SUVR values of target VOIs**

|  | HC | AD | PSP |
| --- | --- | --- | --- |
| Braak Ⅴ/Ⅵ SUVR |  |  |  |
| Quantified by CB-ref | 0.83 (0.08) | 1.48 (0.31) | 0.86 (0.11) |
| Quantified by GM-ref | 0.88 (0.05) | 1.41 (0.33) | 0.93 (0.07) |
| Quantified by WM-ref | 0.89 (0.12) | 1.43 (0.24) | 0.94 (0.10) |
| STN SUVR |  |  |  |
| Quantified by CB-ref | 1.28 (0.18) | 1.32 (0.28) | 1.90 (0.45) |
| Quantified by GM-ref | 1.36 (0.20) | 1.25 (0.27) | 2.08 (0.55) |
| Quantified by WM-ref | 1.37 (0.25) | 1.26 (0.17) | 2.06 (0.44) |

HC, healthy control; AD, Alzheimer’s disease; PSP, progressive supranuclear palsy; SUVR, standardized uptake value ratio; VOI, volume of interest; Braak Ⅴ/Ⅵ, composite areas consisting of neocortex; STN, subthalamic nucleus; CB-ref, cerebellar reference; GM-ref, gray matter reference; WM-ref, white matter reference

Values are listed as mean ± standard deviation
